# The correlations of outcomes and vascular morphology with infarct patterns in middle cerebral arterial trunk occlusion

**DOI:** 10.1101/2025.01.10.25320365

**Authors:** ZhiRong Cai, Yuan Chen, ShaoQing Pei, Yue He, YaNan Zhu, Rui Zhang, JingWei Lin, Yi Yang, Ying Zhu

## Abstract

**Background and purpose:** The large vessel occlusion (LVO) in middle cerebral artery (MCA) trunk (i.e., M_1_ segment) caused by intracranial atherosclerotic disease (ICAD) could introduce with different infarct patterns. We aimed to compare the clinical outcomes among these patterns and investigate the associations between the morphological parameters of contralateral MCA (cMCA) M_1_ segment and infarct patterns.

**Methods:** Patients with stroke attributed to M_1_-ICAD-LVO were enrolled. The infarct patterns were categorized into artery-artery embolism (AAE), large infarct, borderzone infarct (BZI) and perforating artery infarction (PAI). The morphological parameters of cMCA-M_1_ segment consisted of proximal diameter, distal diameter, arc length and chord length were measured. The tortuosity index of cMCA-M_1_ segment was calculated by (arc length/chord length-1) × 100%.

**Results:** A total of 171 subjects were enrolled. Compared to AAE, the risk of poor outcome significantly increased in BZI (odds ratio [OR]= 5.51, 95% confidence interval [CI] = 1.71–17.78, *p* = 0.004) and large infarct (OR= 10.92, 95% CI = 2.01–59.27, *p* = 0.006), and was comparable in PAI. The tortuosity index (OR= 2.85, 95% CI = 1.13–7.18, *p* = 0.026) and arc length (OR= 2.47, 95% CI = 1.02–5.97, *p* = 0.045) significantly elevated in BZI and were identical in other three patterns. Subjects other than BZI were categorized into large infarct (n = 32) and non-large-infarct (n = 46) groups, and the proximal diameter (OR= 0.22, 95% CI = 0.07–0.72, *p* = 0.012), arc length (OR= 0.88, 95% CI = 0.78–0.98, *p* = 0.018) and chord length (OR= 0.87, 95% CI = 0.77–0.995, *p* = 0.042) were all negatively associated with the onset of large infarct.

**Conclusion:** For patients with M_1_-ICAD-LVO, the outcomes of large infarct and BZI were poorer than AAE and PAI. The cMCA-M_1_ segment with elevated tortuosity and arc length was associated with BZI, whereas a thin and short M_1_ segment was correlated with large infarct in patients with a cMCA trunk of low tortuosity.

## Introduction

Approximate 30% patients with ischemic stroke are attributed to large vessel occlusion (LVO), and this subtype of stroke has higher risks of poststroke mortality and disability than other subtypes^1^. Middle cerebral artery (MCA) trunk occlusion, namely M_1_ segment occlusion, takes account for 1/3∼1/2 patients with LVO stroke^2-4^, and intracranial atherosclerotic disease (ICAD) is one of the common pathogenies of LVO^5^, especially in Asian population^6^.

As we observe in clinical practice, ischemic stroke caused by M_1_-ICAD-LVO can be categorized into several patterns: artery-artery embolism (AAE), large infarct, border-zone infarct (BZI), perforating artery infarction (PAI) and mixed pattern, in line with some previous works^7-9^. Why are infarct patterns derived from ICAD-LVO heterogeneous rather than a uniform pattern with a large ischemic lesion within the corresponding territory? This maybe due to the different extents of collaterals between individuals^10^. In case of occlusion in a major supply artery, the blood flow in the collaterals will be redistributed by the cerebral automatic regulation mechanism to sustain the supply for the ischemic core and peri-infarct region^11^. This compensated mechanism may contributes to provide partial flow to the territory of occlusive MCA thus decrease the ischemic volume^12^, and ultimately represents with different infarct patterns.

The poststroke outcomes of these infarct patterns are with large disparities. The rate of poor clinical outcome is higher than 70% in patients with a large infarct even though with endovascular treatments ^13-15^, whereas is approximately 20% in patients with BZI or PAI^16, 17^. However, the comparisons of poor outcomes among these patterns in the context of M_1_-ICAD-LVO are still unknown. Exploring the pathogenesis underlying the infarct patterns caused by M_1_-ICAD-LVO is needed because of the probable diversity of their clinical outcomes. According to previous reports, AAE is likely to be involved with the presence of vulnerable atherosclerotic plaque^18^; a large infarct maybe introduced by a complete occlusion resulted from in situ thrombosis^7^; BZI usually indicates insufficient blood flow in culprit vessel^19^; and PAI is associated with the blockage of the orifice of the perforating artery by a parental arterial atheroma^20, 21^. In a word, the pathogenesis probably correlates with impaired hemodynamics in BZI and with the atherosclerotic plaque features of MCA in other three infarct patterns.

The vascular tortuosity can mediate the progress of atherosclerotic plaque through the effect on the hemodynamics^22^, and a study has demonstrated the vascular tortuosity is associated with the occurence of plaque in MCA^23^. A recent study based on the high-resolution vessel wall imaging has further found the vascular tortuosity involves with the plaque features^24^. In addition, the increased vascular tortuosity results in local turbulence in the vessels thereby impairs the territorial hemodynamics^25^. Other than the vascular tortuosity, the luminal diameter can promote the plaque rupture and thrombosis^26^ through the impact on the plaque structural stress^27^. In summary, vascular morphology seems to have an impact on the luminal plaque features and hemodynamics, thereby potentially associating with the infarct patterns caused by M_1_-ICAD-LVO.

This study compares the clinical outcomes of various infarct patterns attributed to M_1_-ICAD-LVO, and investigates the relationship of these patterns with MCA morphology, aiming to preliminarily understand the clinical outcomes and potential pathogenesis of different infarct patterns and to provide some enlightenment for the clinical managements of M_1_-ICAD patients with a high risk of acute occlusion and subsequent ischemic stroke.

## Methods

### Study populations

This study is a double-center cross-sectional study which conformed to the STROBE guideline and has been approved by local ethics committee. All subjects or their legal guardians have provided the informed consents. We screened all patients with acute or subacute ischemic stroke within 14-day of onset who were admitted to the stroke units of Affiliated Hospital of Jiangsu University and Siyang Hospital from 2020.08.01 to 2023.12.31, with the inclusion criteria: 1) ischemic stroke caused by M_1_-ICAD-LVO; 2) with an age of 18-year or older. Patients with potential cardioembolic source (e.g., atrial fibrillation, cardiomyopathy, patent foramen ovale); with intravenous thrombolysis and/or endovascular treatment; with M_1_-LVO due to non-ICAD (e.g., dissection, vasculitis, and Moyamoya disease); with poor neuroimaging quality or lack of neuroimaging evidence; with occlusion in bilateral MCAs or anterior cerebral arteries (ACAs); with tandem stenosis in ipsilateral carotid artery; with a large chronic infarct in the territory of occlusive MCA; with functional dependence before the index stroke (i.e., modified Ranking Scale [mRS] 3∼5 points) were excluded. PAI resulted from the blockage of the orifice of the perforating artery by ICAD in parental artery usually involves with the lowest section of the basal ganglia^28^, thus those without the infarct extending to the basal ganglia were also excluded.

On admission, all clinical data of subjects, including age, sex, a history of hypertension and diabetes mellitus, smoking and a history of previous stroke (including both ischemic and hemorrhagic stroke), were collected by a face to face interview. The severity of neurological deficit of subjects were evaluated by a neurologist via National Institute of Health Stroke Scale (NIHSS), and all subjects were administrated according to the up-to-date guidelines^29, 30^. During the hospitalization, all subjects underwent the neuroimaging and laboratory tests, and tests of the cardiac diseases, such as 24-hour dynamic electrocardiogram and cardiac ultrasound, etc. The daily activities of all subjects were evaluated according to mRS (0∼5 points, death was recorded as 6 points) at discharge. A poor clinical outcome referred to a mRS of 3∼6 points at discharge.

### The infarct patterns evaluated via neuroimaging

The evaluations of the infarct patterns were performed by using CT scan or diffused weighted image of magnetic resonance imaging. The infarct patterns were categorized based on Chinese Ischemic Stroke Subclassification^31^ as follows: 1) AAE: single or multiple small infarcts limited to the cortex, with or without small subcortical infarct(s). A large wedge- or oval-shaped infarct limited to the cortex in the territory of MCA but not involved with the border-zone was also categorized as AAE^8^ (Figure 1: A1 and A2); 2) a large infarct: the volume of the ischemic lesion involved with both cortical and subcortical regions was 70 ml or greater in CT scan or diffused weighted image^15^ (Figure 1: B); 3) BZI: this pattern was divided into external cortical border-zone and internal border-zone infarcts. The former referred to the linear- or wedge- or oval-shaped lesion(s) in the regions between the cortical territories of ACA and MCA or between MCA and posterior cerebral artery, the latter was defined as infarcts in the white matter along and (or) above the lateral ventricles between the deep and the superficial supplying territories of MCA or between the superficial territories of MCA and ACA, including confluent and partial subtypes. The confluent internal BZI represented as a large cigar shape, the partial internal BZI as a linear-shaped lesion paralleling to the lateral ventricle with at least 3 lesions and each lesion with a diameter of 3 mm or larger^7, 8^ (Figure 1: C1 to C4); 4) PAI: single or multiple subcortical infarcts extending from the lowest portion of basal ganglia to radiating corona or semioval center^8, 28^. In case of multiple subcortical infarcts, the distribution of the lesion differed from the the partial internal BZI (Figure 1: D1 to D6).

**Figure 1.**
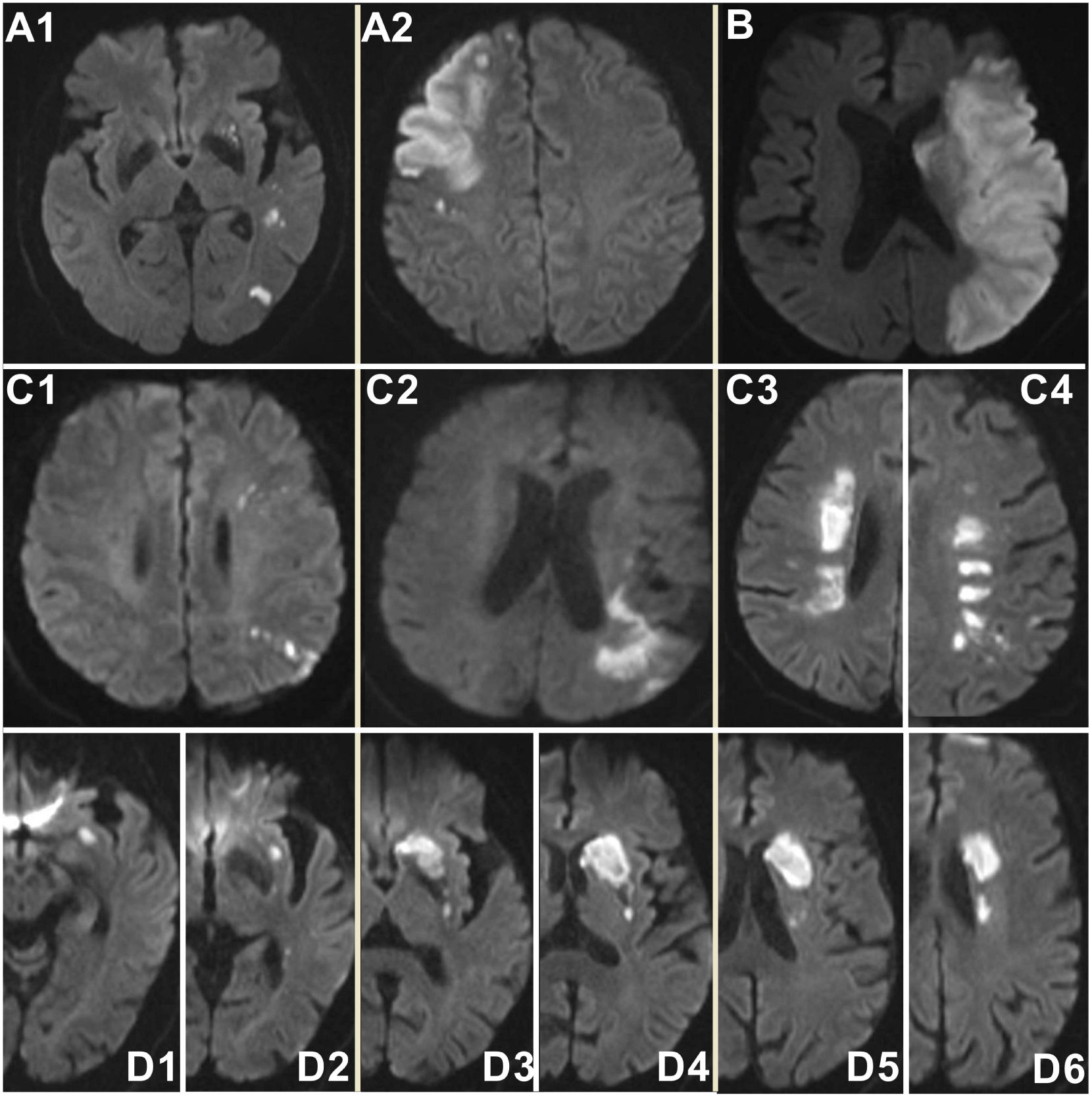
Graphical representations of various infarct patterns caused by M_1_-ICAD-LVO. (A1 and A2): artery-artery embolism; (B): large infarct; (C1 and C2): the cortical external border-zone infarcts; (C3): the confluent internal border-zone infarcts; (C4): the partial internal border-zone infarcts; (D1 to D6): perforating artery infarction. Abbreviations: ICAD indicates intracranial atherosclerotic disease; LVO, large vessel occlusion.

Some scholars suggested the infarct patterns caused by MCA-ICAD included a mixed pattern^8, 9^. The mixed patterns of the subjects in this study represented in manner of BZI combined with AAE or PAI. The hypoperfusion could impair the clearance of emboli which would block the cortical or perforating arteries thus result in AAE or PAI^32^. Therefore, we considered that the hypoperfusion played an important role in the mixed patterns, and these patterns were also categorized as BZI in this study, in accordance with a previous work^33^.

Two independent neurologists evaluated the infarct patterns respectively and a superior practitioner made the final diagnosis in case of disagreement between the two evaluators.

### The MCA morphology measured via magnetic resonance or CT angiography

The morphology of the culprit MCA could not be measured due to the complete occlusion. A previous study reported the morphological features of bilateral MCAs were with no significant differences in humans^23^, we therefore measured the morphology of contralateral MCA (cMCA) to surrogate that of the culprit MCA.

The method of the measurement of the cMCA-M_1_ segment morphology in this study was in line with a previous work^23^. MCA-M_1_ segment referred to the vessel from the bifurcation of ACA and MCA to the MCA bifurcation. We measured the morphological parameters of cMCA-M_1_ segment, including the proximal diameter, the distal diameter, the arc length and the chord length.

The shape of cMCA-M_1_ segment mainly consisted of U, S and linear shapes^34^. The morphological parameters of linear-shaped cMCA were measured in any section with clearly exposed cMCA-M_1_ segment chosen from the forward 3-dimensional cerebrovascular imaging in magnetic resonance or CT angiography. For U- and S-shaped cMCA-M_1_ segment, we rotated the forward 3-dimensional cerebrovascular imaging along the horizontal axis and selected the section with the smallest included angle of cMCA-M_1_ segment to measure the morphological parameters. Two lines starting respectively from the ACA-MCA bifurcation and MCA bifurcation and running along the central line of the vessel were drawn, and the angle at the point where these two lines met was the included angle of U-shaped cMCA-M_1_ segment (Figure 2).

**Figure 2.**
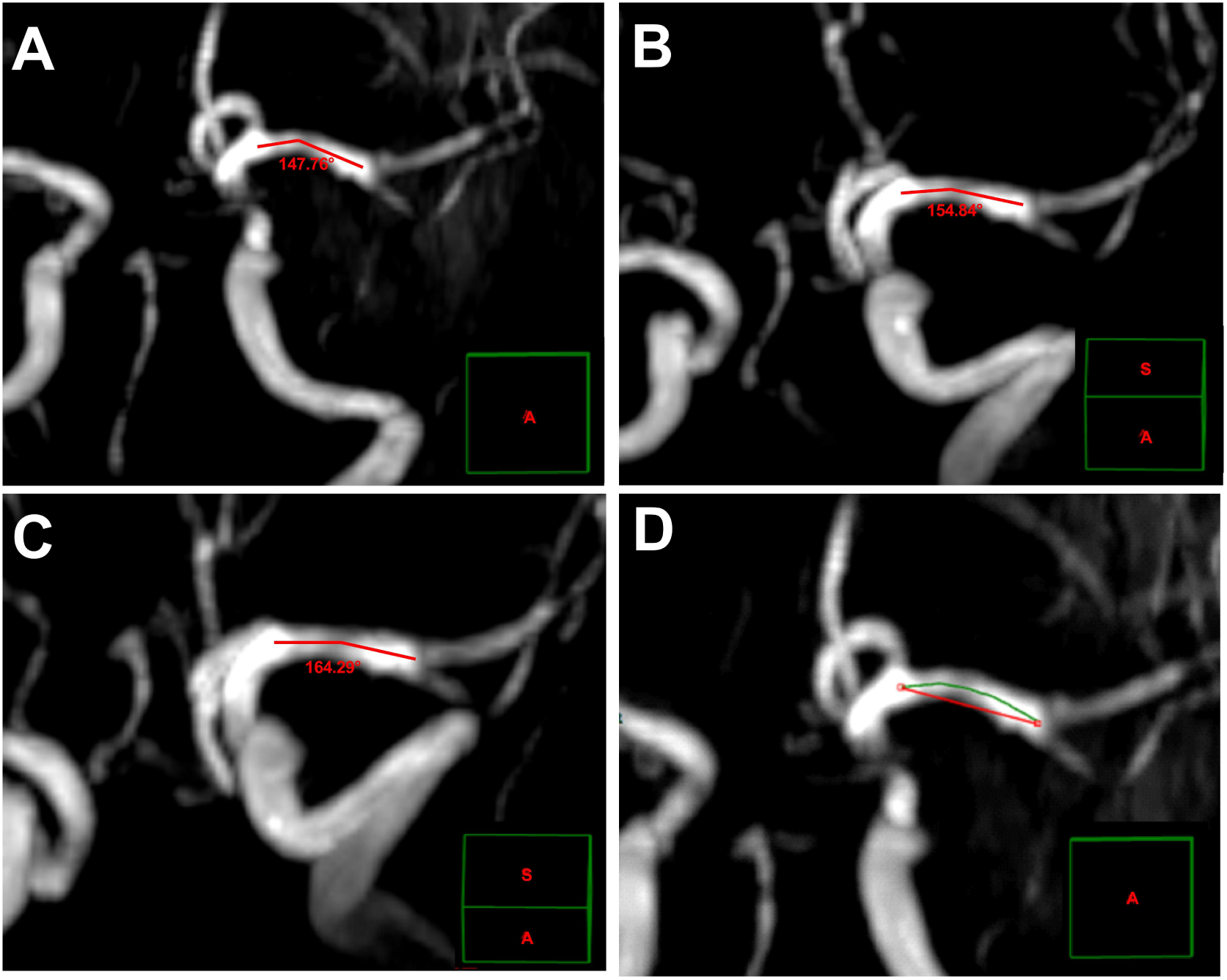
The measurements of the morphological parameters in U-shaped cMCA. (A to C): the measurements of included angle (at the point where the two red lines met) of cMCA trunk; (D): the section with the smallest included angle (A) was used to measure the arc length (green line) and the chord length (red line). The green line box in the lower right corner of each image indicates the spatial position of this section in the forward 3-dimensional cerebrovascular imaging. Abbreviations: cMCA indicates contralateral middle cerebral artery.

For S-shaped cMCA-M_1_ segmen, the included angles of two major turning points were measured. One angle was between the central line starting from ACA-MCA bifurcation and the central line of the mainstream of the middle part of the vessel (Angle a), whereas the latter and the central line starting from the MCA bifurcation formed the other angle (Angle b) (Figure 3). The section with the smallest sum of the two angles were selected. If the contralateral ACA-A_1_ segment was lack, we estimated the approximate starting point of cMCA according to the site of ipsilateral ACA-MCA bifurcation.

**Figure 3.**
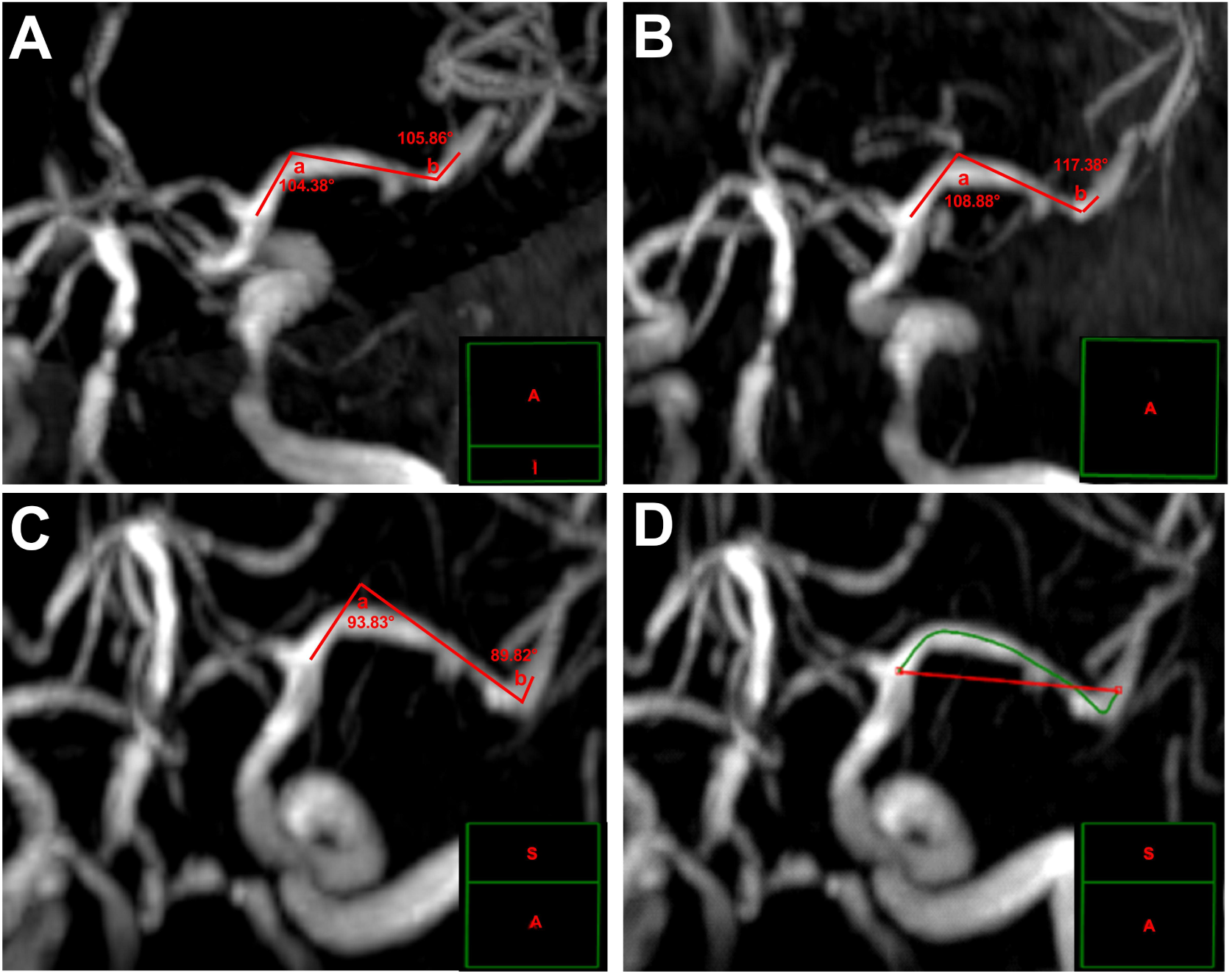
The measurements of the morphological parameters in S-shaped cMCA. (A to C): the measurements of included angle (Angle a and b) of cMCA trunk; (D): the section with the smallest sum of included angles (C) was used to measure the arc length (green line) and the chord length (red line). The green line box in the lower right corner of each image indicates the spatial position of the this section in the forward 3-dimensional cerebrovascular imaging. Abbreviations: cMCA indicates contralateral middle cerebral artery.

The arc length was the length of the arc lying between the ACA-MCA bifurcation and the MCA bifurcation along the central line of the trunk, and the chord length referred to the minimal distance between the two bifurcations. The formula of the tortuosity index of cMCA was (arc length/chord length-1) × 100%^35^. The proximal and distal diameters of cMCA were measured within 2 mm from the two bifurcations, perpendicular to the direction of the trunk.

The work software Carestream RISGC (Carestream Health, USA, version 3.1.S05.0) was used to measure the morphological parameters of cMCA. Two neurologists (Y He and R Zhang) blinded to the clinical datum of all subjects respectively measured the morphological parameters of cMCA-M_1_ segment, the mean of which were used in the statistical analyses.

We also evaluated whether cMCA-M_1_ segment had ICAD which was reported to be associated with the vascular morphology^23^. ICAD referred to any degree of atherosclerotic stenosis in cMCA-M_1_ segment. The stenotic degree was evaluated according to Warfarin-Aspirin Symptomatic Intracranial Disease Study Trial method.

### Reproducibilities in the infarct pattern evaluation and cMCA morphology measurements

To assess the intra- and inter-observer agreements, two neurologists with working experience of 10-year or more (Z.R. Cai and Y. Chen) evaluated the infarct patterns of all subjects twice, respectively, with at least 1-week interval between two evaluations for each subject. Kappa test was used in the assessments of intra- and inter-observer agreements in the evaluations of infarct pattern, and Bland-Altman test was used in the assessments of inter-observer agreements in the measurements of the cMCA-M_1_ morphological parameters.

### Statistical methods

Software SPSS (version 25.0) was used to perform the statistical analyses. At first, we completed the missing data by using multiple imputation (5 imputations) and selected the set of data with the highest Cronbach’s alpha coefficient in reliability analysis for statistical analysis. Normally distributed continuous variables were described by the mean ± standard deviation (SD) and non-normally distributed continuous variables were described by the median (inter-quartile range, IQR). The comparisons of categorical variables were performed by using chi-squared test or Fisher’s exact test. Kruskal-Wallis test or one-way analysis of variance test was used to compare the continuous variables among multiple groups, and independent sample *t* test or Wilcoxon test was used to compare the continuous variables between two groups.

The subjects were divided into four groups based on the infarct patterns subclassification and univariate analyses among these groups were performed. Age, sex and confounders with a *p* value < 0.2 were included into multivariable regression model. The binary logistic regression was used to analyze the association between the infarct pattern and poor clinical outcomes. According to the tertiles of the vascular morphological parameters with a significant difference in the univariable analyses, we trichotomized the subjects respectively and analyzed the associations between the infarct patterns and the levels of these parameters in a ordinal logistic regression model. Due to the lowest rate of poor outcome, AAE group was used as the reference in multivariable analyses. The receiver operating characteristic (ROC) curve was used to analyze the correlation of BZI and the vascular morphological parameters. Consequently, we divided the subjects other than BZI patients into large infarct and non-large-infarct groups, and the binary logistic regression and ROC curve was used to analyze the association between the morphological parameters and the onset of large infarct. All tests were two sided and a *p* value of < 0.05 was considered statistically significant.

## Results

A total of 309 patients with acute or subacute ischemic stroke attributed to M_1_-LVO were screened. Among these patients, 50 with potential cardioembolic source, 42 with intravenous thrombolysis and (or) endovascular treatment, 4 with suspicious Moyamoya disease, 10 with poor neuroimaging quality or without neuroimaging evidence, 12 with bilateral MCA or ACA occlusion, 8 with tandem stenosis in ipsilateral carotid artery, 4 with a chronic large infarct within the territory of MCA and 8 with PAI not involving with the lowest section of basal ganglia, these patients were all excluded. Finally, 171 subjects were enrolled in this study, 98 from Affiliated Hospital of Jiangsu University and 73 from Siyang Hospital ( Figure 4).

**Figure 4.**
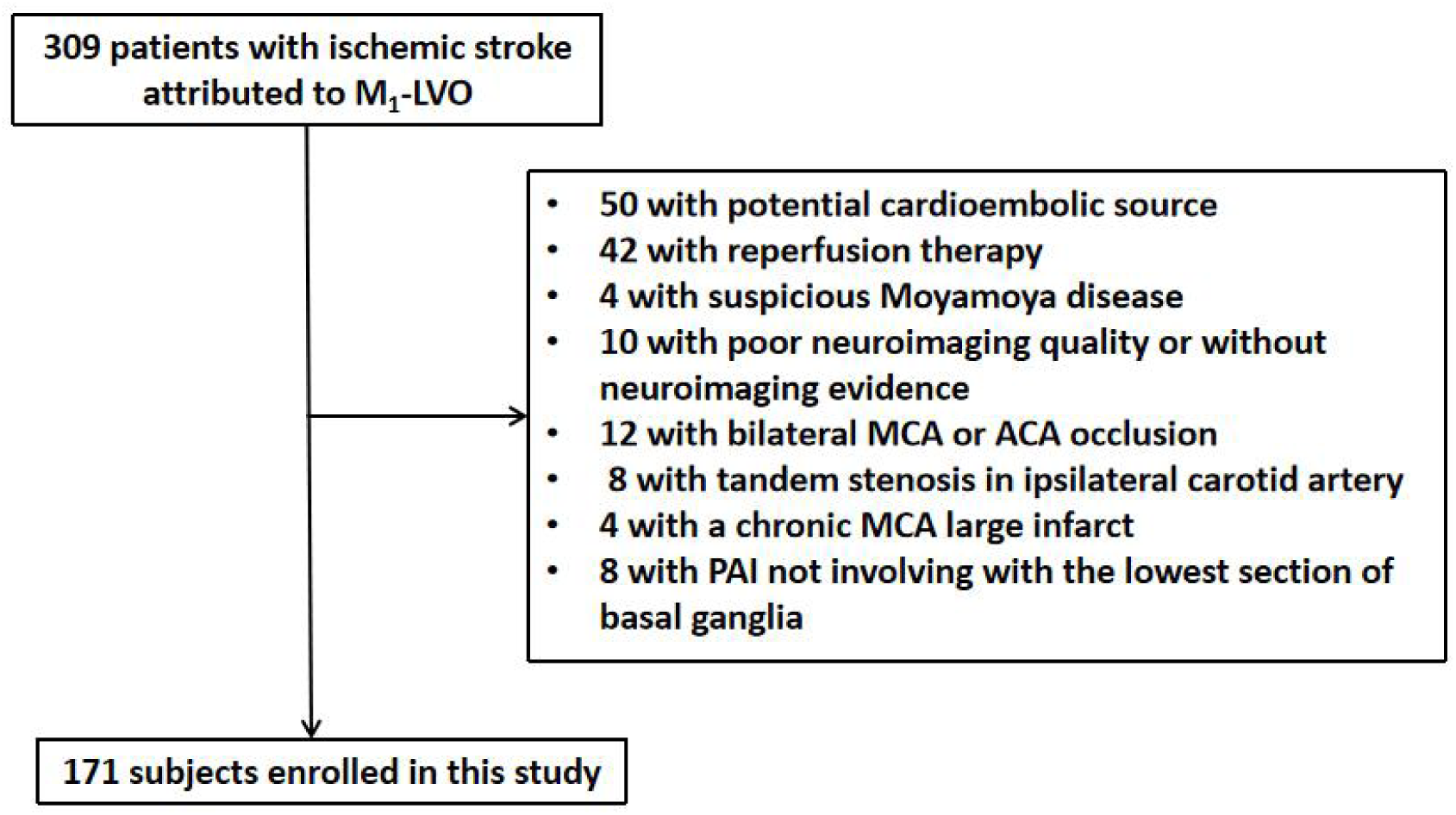
Flow diagram of subjects’ enrollment. Abbreviations: LVO indicates large vessel occlusion; MCA, middle cerebral artery; ACA, anterior cerebral artery; PAI, perforating artery infarction.

### Baseline data of subjects

The median age of 171 subjects were 71.0 years (62.0,77.0 years), and 95 (55.6%) patients were male. The median of initial NIHSS was 4.0 points (2.0, 6.0 points). Of all subjects, 31 (19.9%) were with subclassification of AAE, 32 (20.5%) with large infarct, 93 (59.6%) with BZI and 15 (8.8%) with PAI. 28 (16.4%) patients had ICAD in cMCA. The morphological parameters of all subjects were as follows: with the mean proximal diameter of 2.58 mm (with a SD of 0.61 mm), with the median of the distal diameter of 2.08 mm (1.74, 2.45 mm) and with the medians of arc length and chord length of 22.73 mm (18.91, 27.06 mm) and 19.66 mm (16.42, 23.46 mm), respectively. The tortuosity index of cMCA had a median of 14.95% (7.97, 22.29 %). 91 (53.2%) patients suffered from a poor clinical outcome at discharge (Supplementary Table S1).

### The reproducibilities of infarct pattern evaluation and cMCA morphology measurements

The intra-observer agreements (with κ values of 0.916 and 0.907, respectively, and with both *p* values of < 0.001) and the inter-observer agreements (with κ value of 0.860 and with a *p* value of < 0.001) of the infarct pattern evaluation were excellent. For the measurements of vascular morphological parameters, the inter-observer agreements were acceptable ( 97.7% of the proximal diameter values, 97.1% of distal parameter and 94.7% of tortuosity index were within 95% limit of agreement in Bland-Altman test) (Supplementary Figure S1).

### The comparison of clinical outcomes among four infarct patterns

Initial NIHSS (*p* < 0.001), the arc length (*p* < 0.001), chord length (*p* = 0.005) and tortuosity index (*p* = 0.002) of cMCA and the rate of poor outcome (*p* < 0.001) were significantly different among the four patterns in univariable analyses (Table 1). In the binary logistic regression model, after adjusting for age, sex and factors with a *p* < 0.2 in univariable analyses, including a history of previous stroke, diastolic blood pressure at admission, levels of total cholesterol and low-density lipoprotein cholesterol, initial NIHSS, proximal diameter and tortuosity index of cMCA, the risks of poor outcome in BZI (odds ratio [OR]= 5.51, 95% confidence interval [CI] = 1.71–17.78, *p* = 0.004) and large infarct (OR= 10.92, 95% CI = 2.01–59.27, *p* = 0.006) groups were higher than in AAE group. The PAI group had a comparable risk of poor outcome than the AAE group (Table 2).

**Table 1.** The univariable analyses of clinical characteristics among different infarct patterns.

| Clinical characteristics | AAE<br>(n=31) | Large infarct<br>(n=32) | PAI<br>(n=15) | BZI<br>(n=93) | P-value |
| --- | --- | --- | --- | --- | --- |
| Male, n (%) | 17 (54.8) | 18 (56.3) | 11 (73.3) | 49 (52.7) | 0.52 |
| Age (year), median (IQR) | 71.0 (63.0, 80.0) | 74.5 (68.0, 79.0) | 69.0 (62.0, 77.0) | 69.0 (62.0, 77.0) | 0.067 |
| Hypertension, n (%) | 22 (71.0) | 25 (78.1) | 13 (86.7) | 68 (73.1) | 0.67 |
| Diabetes mellitus, n (%) | 8 (25.8) | 6 (18.8) | 2 (13.3) | 25 (26.9) | 0.65 |
| Smoking, n (%) | 13 (41.9) | 9 (28.1) | 8 (53.3) | 33 (35.5) | 0.36 |
| Previous stroke, n (%) | 14 (45.2) | 7 (21.9) | 3 (20.0) | 24 (25.8) | 0.15 |
| SBP (mmHg), mean ± SD | 147.7 ± 22.1 | 149.6 ± 34.0 | 159.9 ± 23.9 | 150.4 ± 18.5 | 0.40 |
| DBP (mmHg), mean ± SD | 82.0 ± 12.0 | 81.8 ± 13.6 | 89.9 ± 10.8 | 83.6 ± 12.6 | 0.18 |
| TC (mmol/L), mean ± SD | 4.38 ± 0.99 | 4.94 ± 1.17 | 4.46 ± 1.04 | 4.82 ± 1.20 | 0.15 |
| TG (mmol/L), median (IQR) | 1.35 (0.85, 2.24) | 1.27 (1.02, 1.59) | 1.41 (0.96, 1.71) | 1.34 (0.98, 1.87) | 0.87 |
| LDL-C (mmol/L), mean ± SD | 2.41 ± 0.69 | 2.90 ± 0.96 | 2.50 ± 0.94 | 2.72 ± 0.91 | 0.13 |
| HbA1c (%), median (IQR) | 6.30 (5.70, 7.64) | 6.25 (5.50, 7.88) | 5.80 (5.50, 6.80) | 6.00 (5.55, 7.05) | 0.53 |
| Hcy (mmol/L), median (IQR) | 15.20 (10.80, 19.49) | 11.87 (8.34, 17.10) | 14.13 (12.60, 17.50) | 12.40 (8.30, 17.68) | 0.24 |
| Platelet count ($\times 10^9/L$), mean ± SD | 217.0 ± 68.7 | 195.8 ± 63.9 | 188.0 ± 47.7 | 212.4 ± 58.5 | 0.26 |
| cMCA morphology |  |  |  |  |  |
| Proximal diameter (mm),<br>mean ± SD | 2.64 ± 0.57 | 2.43 ± 0.45 | 2.82 ± 0.52 | 2.56 ± 0.66 | 0.19 |
| Distal diameter (mm),<br>median (IQR) | 2.30 (1.66, 2.46) | 2.07 (1.75, 2.51) | 2.33 (2.03, 2.51) | 2.00 (1.67, 2.42) | 0.27 |
| Proximal to Distal diameter ratio,<br>median (IQR) | 1.28 (1.05, 1.43) | 1.13 (0.96, 1.34) | 1.22 (1.14, 1.34) | 1.24 (1.09, 1.54) | 0.20 |
| Arc length (mm), median (IQR) | 21.66 (16.68, 26.24) | 18.59 (16.13, 23.18) | 25.38 (21.41, 27.93) | 25.25 (20.59, 29.56) | <0.001* |
| Chord length (mm), median (IQR) | 19.01 (15.46, 22.17) | 17.68 (13.62, 20.51) | 19.78 (18.05, 25.32) | 20.61 (17.03, 24.43) | 0.005* |
| Tortuosity index (%), median (IQR) | 9.26 (6.75, 18.81) | 10.43 (5.49, 18.96) | 12.87 (6.57, 22.09) | 17.45 (10.28, 30.51) | 0.002* |
| ICAD in cMCA, n (%) | 5 (16.1) | 4 (12.5) | 1 (6.7) | 18 (19.4) | 0.67 |
| Initial NIHSS (point), median (IQR) | 2.0 (1.0, 4.0) | 9.0 (5.0, 13.0) | 3.0 (1.0, 4.0) | 3.0 (2.0, 5.0) | <0.001* |
| Poor outcome, n (%) | 6 (19.4) | 29 (90.6) | 4 (26.7) | 52 (55.9) | <0.001* |

Abbreviations: AAE indicates artery-artery embolism; PAI, perforating artery infarction; BZI, borderzone infarct; SBP, systolic blood pressure; DBP, diastolic blood pressure; TC, total cholesterol; TG, triglyceride; LDL-C, low-density lipoprotein cholesterol; HbA1c, glycosylated hemoglobin; Hcy, homocysteine; cMCA, contralateral middle cerebral artery; ICAD, intracranial atherosclerotic disease; NIHSS, National Institutes of Health Stroke Scale.
\* $p < 0.05$ was considered statistically significant.

**Table 2.** The association between the infarct patterns and poor outcome.

| Infarct pattern | Poor outcome |  |  |  |
| --- | --- | --- | --- | --- |
|  | Model 1<sup>†</sup> |  | Model 2<sup>‡</sup> |  |
|  | Adjusted OR | P-value | Adjusted OR | P-value |
| AAE | Ref. | / | Ref. | / |
| PAI | 1.26 (0.25–6.32) | 0.78 | 1.42 (0.28–7.26) | 0.68 |
| BZI | 4.83 (1.54–15.10) | 0.007* | 5.51 (1.71–17.78) | 0.004* |
| Large infarct | 11.33 (2.09–61.36) | 0.005* | 10.92 (2.01–59.27) | 0.006* |

<sup>†</sup> Adjusting for common factors, including age, sex, previous stroke, diastolic blood pressure at admission, the levels of total cholesterol and low-density lipoprotein cholesterol and initial NIHSS.
<sup>‡</sup> Adjusting for common factors plus proximal diameter and tortuosity index of cMCA trunk.
Abbreviations: AAE indicates artery-artery embolism; PAI, perforating artery infarction; BZI, borderzone infarct.
\* $p < 0.05$ was considered statistically significant.

### The associations between the infarct patterns and the vascular morphological parameters

In the univariable analyses, three parameters including the arc length, the chord length and the tortuosity index of cMCA-M_1_ segment were significantly different among the four groups. According to the tertiles of these three parameters, we trichotomized all subjects respectively. Using the three levels of these parameters as dependent variables in the ordinal logistic regression, respectively, after adjusting for age, sex, a history of previous stroke, diastolic blood pressure at admission, levels of total cholesterol and low-density lipoprotein cholesterol, initial NIHSS and the proximal diameter of cMCA-M_1_ segment, the incidences of elevated arc length (OR= 2.47, 95% CI = 1.02–5.97, *p* = 0.045) and tortuosity index (OR= 2.85, 95% CI = 1.13–7.18, *p* = 0.026) of cMCA-M_1_ segment in BZI groups are higher than in AAE group, whereas these two parameters of PAI and large infarct groups were with no difference compared to AAE group. The chord length among these four groups were comparable (Table 3).

**Table 3.** The associations between the infarct patterns and the morphological parameters of cMCA.

| Infarct pattern | Elevated level of cMCA morphological parameters |  |  |  |  |  |
| --- | --- | --- | --- | --- | --- | --- |
|  | Arc length |  | Chord length |  | Tortuosity index |  |
|  | Adjusted OR<sup>†</sup> | P | Adjusted OR<sup>†</sup> | P | Adjusted OR<sup>†</sup> | P |
| AAE | Ref. | / | Ref. | / | Ref. | / |
| Large infarct | 0.66 (0.21–2.12) | 0.49 | 0.66 (0.21–2.08) | 0.48 | 1.30 (0.36–4.71) | 0.69 |
| PAI | 2.45 (0.84–7.28) | 0.099 | 1.57 (0.58–4.26) | 0.38 | 2.03 (0.60–6.90) | 0.26 |
| BZI | 2.47 (1.02–5.97) | 0.045* | 1.63 (0.71–3.74) | 0.25 | 2.85 (1.13–7.18) | 0.026* |

† Adjusting for age, sex, previous stroke, diastolic blood pressure at admission, total cholesterol, low-density lipoprotein cholesterol, initial NIHSS and proximal diameter of cMCA.
Abbreviations: cMCA indicates contralateral middle cerebral artery; AAE, artery-artery embolism; PAI, perforating artery infarction; BZI, borderzone infarct.
\* $p < 0.05$ was considered statistically significant.

### The associations between the cMCA morphological parameters and onset of large infarct

Because that two distinctive cMCA-M_1_ morphological parameters of BZI had been identified, this category of patients were excluded. The remaining 78 subjects were dichotomized into non-large-infarct (n = 46) and large infarct (n = 32) groups. The levels of total cholesterol (*p* = 0.033) and low-density lipoprotein cholesterol (*p* = 0.021), the proximal diameter (*p* = 0.027), arc length (*p* = 0.009) and chord length (*p* = 0.025) of cMCA-M_1_ segment were significantly different and the tortuosity index was comparable between the two groups (Supplementary Table S2).

After adjusting for age, sex and factors with a *p* < 0.2 in univariable analyses including levels of total cholesterol, low-density lipoprotein cholesterol and homocysteine, the proximal diameter (OR= 0.22, 95% CI = 0.07–0.72, *p* = 0.012), arc length (OR= 0.88, 95% CI = 0.78–0.98, *p* = 0.018) and chord length (OR= 0.87, 95% CI = 0.77–0.995, *p* = 0.042) of cMCA-M_1_ segment were all negatively associated with the onset of large infarct (Table 4).

**Table 4.** The associations between the morphological parameters of cMCA and the onset of large infarct.

| cMCA morphology | Large infarct |  |  |  |
| --- | --- | --- | --- | --- |
|  | Crude OR | P-value | Adjusted OR<sup>†</sup> | P-value |
| Proximal diameter | 0.35 (0.13–0.91) | 0.032* | 0.22 (0.07–0.72) | 0.012* |
| Arc length | 0.89 (0.82–0.98) | 0.016* | 0.88 (0.78–0.98) | 0.018* |
| Chord length | 0.89 (0.79–0.99) | 0.030* | 0.87 (0.77–0.995) | 0.042* |

<sup>†</sup> Adjusting for age, sex, total cholesterol, low-density lipoprotein cholesterol and homocysteine.
Abbreviations: cMCA indicates contralateral middle cerebral artery.
\* $p < 0.05$ was considered statistically significant.

The cMCA-M_1_ morphological parameters between AAE and PAI groups were with no differences (Supplementary Table S3).

### ROC analyses

The area under the curve (AUC) in the indicative value of the cMCA-M_1_ arc length for the onset of BZI was 0.660 (95%CI: 0.579–0.741, *p* < 0.001), with a cut-off value of 26.86 mm, of which the sensitivity was 39.8% and the specificity was 89.7%. The AUC in the tortuosity index indicative of BZI was 0.664 (95%CI: 0.583–0.744, *p* < 0.001), with a cut-off value of 14.65%, and the sensitivity and specificity were 62.4% and 62.8%, respectively (Figure 5A).

**Figure 5.**
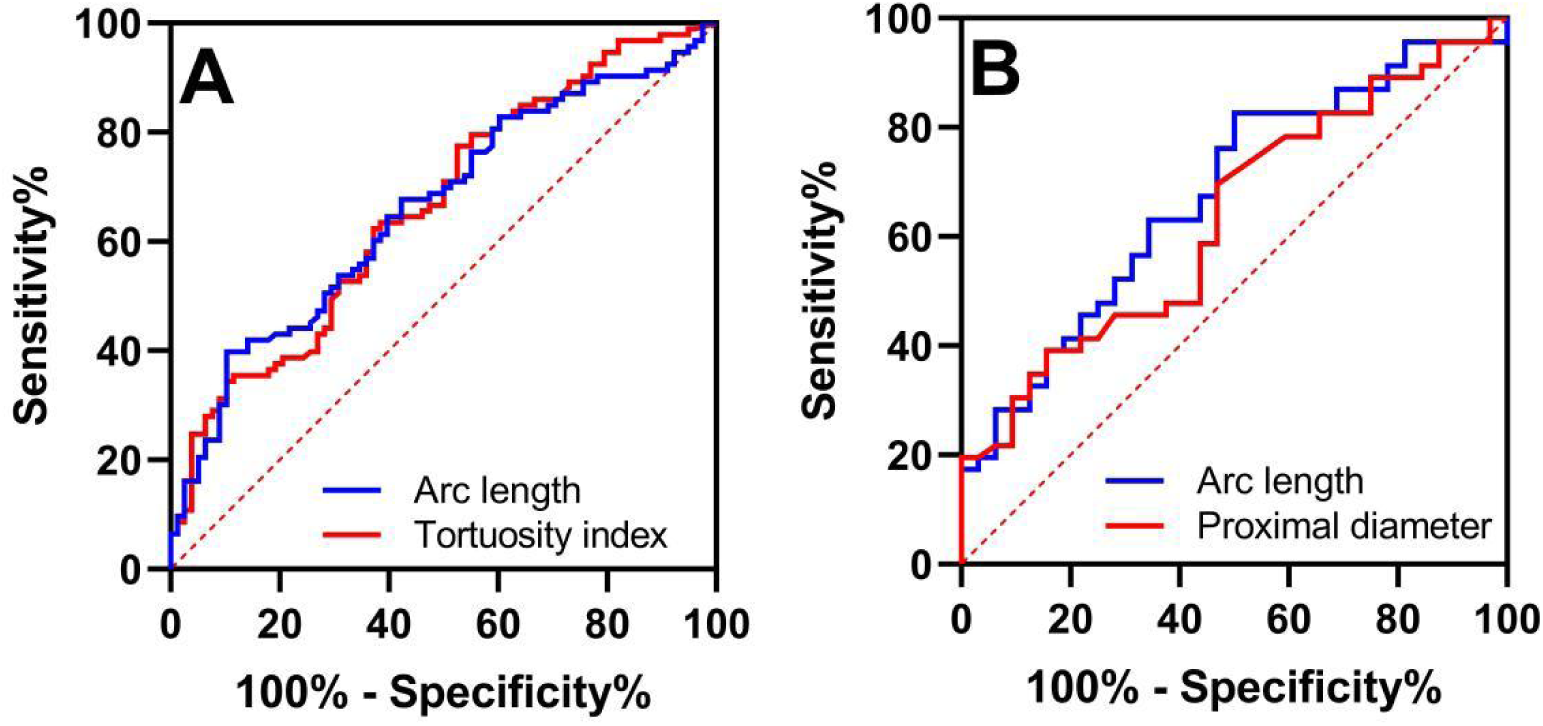
The associations between BZI and morphological parameters and between non-large-infarct and morphological parameters in ROC curve. (A): the AUC of arc length (blue line) was 0.660 (95%CI: 0.579–0.741, *p* < 0.001) and the AUC of tortuosity index (red line) was 0.664 (95%CI: 0.583–0.744, *p* < 0.001) in the indication of BZI; (B): the AUC of arc length (blue line) was 0.676 (95%CI: 0.556–0.796, *p* = 0.009) and the AUC of proximal diameter was 0.633 (95%CI: 0.510–0.757, *p* = 0.046) in the indication of non-large-infarct. Abbreviations: BZI indicates borderzone infarct; ROC, receiver operating characteristic; AUC, area under the curve.

For the indication of the onset of non-large-infarct, the AUC of cMCA-M_1_ proximal diameter was 0.633 (95%CI: 0.510–0.757, *p* = 0.046) and the cut-off value was 2.41 mm, with a sensitivity of 73.9% and a specificity of 50.0%; the AUC of cMCA-M_1_ arc length was 0.676 (95%CI: 0.556–0.796, *p* = 0.009), the cut-off value was 18.32 mm, with a sensitivity of 82.6% and a specificity of 50.0% (Figure 5B).

## Discussion

In this study, we found that in patients with ischemic stroke caused by M_1_-ICAD-LVO, the risk of poor outcomes increased by approximate 4.5-fold and 10-fold in BZI and large infarct compared to AAE, respectively; the arc length and tortuosity index of cMCA-M_1_ segment in BZI were the highest among the four infarct patterns; the proximal diameter, arc and chord length of cMCA-M_1_ segment were negatively associated with the onset of large infarct.

Among the four infarct patterns attributed to the M_1_-ICAD-LVO, the rate of poor outcome of large infarct was the highest and up to 90%, which was in accordance with previous works^13-15^. However, the rate of poor outcome of BZI was 55.9%, far higher than previously reported^16^. The disparity of clinical outcomes between the two studies maybe due to that the stenotic degree of culprit vessel in the present study (all occlusion) is greater than in the previous work (from 50% to occlusion), whereas the severer stenosis of culprit vessel determines a poorer clinical outcome^36^. After adjusting for confounders in the multivariable analyses, the incidence of poor outcome was the the highest in BZI, followed by BZI, and was lowest both in PAI and AAE in this study, suggesting that the risk of poststroke disability of large infarct and BZI patterns was higher than AAE and PAI in the context of MCA trunk occlusion. The extensive loss of eloquent brain matter and substantial poststroke neuroinflammation are the probable reasons of the poor outcome of patients with large infarct^37^; whereas for patients with BZI, the major pathogenesis is hypoperfusion, which could thereafter decrease the blood supply for oligemic tissue surrounding the ischemic penumbra and introduce with the subsequent infarct^38^. These are the potential pathophysiological reasons for the poor outcomes of patients with large infarct or BZI. The relative favorable clinical outcomes of AAE and PAI maybe related to the limited volumes of the ischemic core and penumbra.

Owing to the heterogeneous clinical outcomes of various infarct patterns in the context of M_1_-ICAD-LVO, we explored the vascular morphological features associated with these infarct patterns, in order to roughly understand the pathogenesis underlying these patterns. We found in the univariable analyses that the arc length and tortuosity index of BZI were the largest among the four patterns, which was validated in the further multivariable analyses after adjusting for confounders. This indicates that a long and tortuous MCA trunk seems to correlate with the onset of BZI. As the upstream vessel of the cerebral small vessels, the major intracranial artery with a large tortuosity could not only impair the intraluminal hemodynamics^25^ but also cause the formation of cerebral small vessel disease^39^ and simultaneously affect the blood flow within these small vessels^40^. In addition, the presence of small vessel disease could thereafter result in insufficient collaterals recruitments^41^. The abovementioned pathophysiologies are detrimental to the blood supply for the territory of occlusive MCA, which is the probable mechanism underlying the onset and poor outcome of BZI in patients with M_1_-ICAD-LVO.

The distinctive cMCA-M_1_ morphological parameters of patients with BZI were firstly identified thus these patients were excluded from the further analyses. Because of the poorest clinical outcomes of large infarct among the four patterns, so we attempted to find out the cMCA-M_1_ morphological features of this category of patients. The cMCA morphological parameters were comparable among patients with AAE, PAI and large infarct, we thereafter re-categorized these patients into two groups: large infarct and non-large-infarct groups. Interestingly, subjects of the two groups had significantly different proximal diameters, arc lengths and chord lengths of cMCA-M_1_ segment which were all corroborated to be negatively associated with the onset of large infarct in multivariable regression analyses. This indicates that a ICAD-MCA trunk with a small proximal lumen and a short length is possibly susceptible to the onset of large infarct. A previous work reported that a short vessel was correlated with a large infarct volume^24^, which supported the result of this study, but the mechanism was still unclear. We speculate that a MCA trunk with a thin lumen and a short length is prone to be instantly blocked by rapid growing clots, leading to the formation of large infarct before effective compensation from the collaterals.

The vascular morphological parameters of cMCA-M_1_ segment were identical between AAE and PAI patterns, with a large proximal diameter and a low tortuosity. A MCA trunk with a low tortuosity maybe associated with good ipsilateral collaterals recruitments, and the incidence of large infarct in case of acute occlusion in this trunk is relatively low. However, a large luminal diameter is reported to be correlated with high plaque structural stress^27^ which would cause large necrotic core^42^, upregulated expression of matrix metalloproteinase and macrophage aggregation^43, 44^ in the atherosclerotic plaque. Therefore, the elevated plaque structural stress is likely to be a pivotal factor mediating the plaque rupture, thrombosis and the ultimate vascular events^26^. The onset of AAE maybe attributed to the blockage of the end of cortical small vessel or perforating artery by the debris from vulnerable plaque or embolus. If the orifice of a perforating artery is completely blocked in the process of plaque rupture or thrombosis, the infarct pattern of PAI is probably to occur because that the perforating artery is an end-artery and lack of collaterals^45^.

This study has certain clinical significance for M_1_-ICAD patients who has a high risk of acute occlusion and subsequent ischemic stroke, and could provide preliminary risk stratification for these patients and guide the clinical administrated strategies of subgroups with different potential future infarct patterns. Firstly, M_1_-ICAD patients with a long and a tortuous cMCA-M_1_ segment are susceptible to the onset of BZI and they may not benefit from intensive control of blood pressure^46^. Secondly, for M_1_-ICAD patients with a relatively straight, thin and short cMCA-M_1_ segment, the risk of large infarct is relatively high. A recent study has reported that the balloon angioplasty combined with medical management is more superior than sole medical management for patients with intracranial symptomatic ICAD^47^. Whether endovascular treatment, such as balloon angioplasty, could improve the prognoses of asymptomatic M_1_-ICAD patients who has a high risk of large infarct is needed to be validated in subsequent clinical trails. Lastly, for those with a straight, thick and moderate long cMCA-M_1_ segment, the culprit vessel is susceptible to the formation of vulnerable plaque, which probably leads to the onset of AAE or PAI, thus the intensive statins therapy seems to be important^48^.

This study has some limitations. Firstly, despite as a double-center study, this study has a relatively small sample size and the nature of retrospective study. Whether there is a causal relationship between the cMCA-M_1_ morphological features and the onset of different infarct patterns caused by M_1_-ICAD-LVO needs to be demonstrated in further prospective studies. Secondly, the morphological parameters of the occlusive culprit vessel could not be measured, which was surrogated by the parameters of cMCA. Eventhough that the morphological parameters between the bilateral MCAs were with no difference^23^, the conclusion of this study might be still affected. Thirdly, we excluded all patients with intravenous thrombolysis and endovascular treatment due to the potential effect of reperfusion therapy on the infarct patterns. All subjects enrolled in this study had no indications for reperfusion therapy and the majority of them did not undergo the CT perfusion imaging. Therefore, we did not investigate the association between the cMCA morphological features and the extent of the collaterals for further testifying the pathogenesis underlying different infarct patterns, especially the BZI. Our hypothesis of the pathogenesis of BZI could be partially verified by using CT perfusion imaging in a further study.

## Conclusion

Among the infarct patterns caused by M_1_-ICAD-LVO, the clinical outcome was poorest in large infarct, followed by BZI, and the outcomes of AAE and PAI were relatively favorable. The elevated tortuosity and arc length of cMCA-M_1_ segment were associated with the onset of BZI, whereas small proximal diameter and short arc length were suggestive of the possible onset of large infarct in patients with cMCA of low tortuosity. This study contributed to provide some potential imaging indicators for the future infarct patterns of M_1_-ICAD patients with a high risk of stroke, and also provide guidance for their clinical managements.

## Data Availability

All data generated or analyzed during this study could be acquired from the corresponding author through e-mail.

## References

1. Waqas M, Rai AT, Vakharia K, Chin F, Siddiqui AH. Effect of definition and methods on estimates of prevalence of large vessel occlusion in acute ischemic stroke: a systematic review and meta-analysis. J Neurointerv Surg. 2020;12:260–265

2. Mokin M, Pendurthi A, Ljubimov V, Burgin WS, Siddiqui AH, Levy EI, Primiani CT. Aspects, large vessel occlusion, and time of symptom onset: estimation of eligibility for endovascular therapy. Neurosurgery. 2018;83:122–127

3. Waqas M, Mokin M, Primiani CT, Gong AD, Rai HH, Chin F, et al. Large vessel occlusion in acute ischemic stroke patients: a dual-center estimate based on a broad definition of occlusion site. J Stroke Cerebrovasc Dis. 2020;29:104504

4. Yang S, Wu L, Shi X, Guo C, Yue C, Fan S, et al. Effect of occlusion site on the effectiveness and safety of endovascular thrombectomy for large ischemic cores: a cohort study. Int J Surg. 2024

5. Lee JS, Lee SJ, Hong JM, Alverne F, Lima FO, Nogueira RG. Endovascular treatment of large vessel occlusion strokes due to intracranial atherosclerotic disease. J Stroke. 2022;24:3–20

6. Jia B, Feng L, Liebeskind DS, Huo X, Gao F, Ma N, et al. Mechanical thrombectomy and rescue therapy for intracranial large artery occlusion with underlying atherosclerosis. J Neurointerv Surg. 2018;10:746–750

7. Lee DK, Kim JS, Kwon SU, Yoo SH, Kang DW. Lesion patterns and stroke mechanism in atherosclerotic middle cerebral artery disease: early diffusion-weighted imaging study. Stroke. 2005;36:2583–2588

8. Feng X, Chan KL, Lan L, Abrigo J, Liu J, Fang H, et al. Stroke mechanisms in symptomatic intracranial atherosclerotic disease. Stroke. 2019;50:2692–2699

9. Tekle WG, Hassan AE. Intracranial atherosclerotic disease. Neurology. 2021;97

10. Faber JE, Storz JF, Cheviron ZA, Zhang H. High-altitude rodents have abundant collaterals that protect against tissue injury after cerebral, coronary and peripheral artery occlusion. J Cereb Blood Flow Metab. 2021;41:731–744

11. Binder NF, El Amki M, Glück C, Middleham W, Reuss AM, Bertolo A, et al. Leptomeningeal collaterals regulate reperfusion in ischemic stroke and rescue the brain from futile recanalization. Neuron. 2024;112:1456–1472

12. Elijovich L, Goyal N, Mainali S, Hoit D, Arthur AS, Whitehead M, et al. Cta collateral score predicts infarct volume and clinical outcome after endovascular therapy for acute ischemic stroke: a retrospective chart review. J Neurointerv Surg. 2016;8:559–562

13. Yoshimura S, Sakai N, Yamagami H, Uchida K, Beppu M, Toyoda K, et al. Endovascular therapy for acute stroke with a large ischemic region. N Engl J Med. 2022;386:1303–1313

14. Sarraj A, Hassan AE, Abraham MG, Ortega-Gutierrez S, Kasner SE, Hussain MS, et al. Trial of endovascular thrombectomy for large ischemic strokes. N Engl J Med. 2023;388:1259–1271

15. Huo X, Ma G, Tong X, Zhang X, Pan Y, Nguyen TN, et al. Trial of endovascular therapy for acute ischemic stroke with large infarct. N Engl J Med. 2023;388:1272–1283

16. Yong SW, Bang OY, Lee PH, Li WY. Internal and cortical border-zone infarction: clinical and diffusion-weighted imaging features. Stroke. 2006;37:841–846

17. Yang Y, He Y, Han W, Xu J, Cai Z, Zhao T, et al. Clinical factors associated with functional outcomes in patients with single subcortical infarction with neurological deterioration. Front Neurol. 2023;14:1129503

18. Wu F, Song H, Ma Q, Xiao J, Jiang T, Huang X, et al. Hyperintense plaque on intracranial vessel wall magnetic resonance imaging as a predictor of artery-to-artery embolic infarction. Stroke. 2018;49:905–911

19. Das S, Shu L, Morgan RJ, Shah A, Fayad FH, Goldstein ED, et al. Borderzone infarcts and recurrent cerebrovascular events in symptomatic intracranial arterial stenosis: a systematic review and meta-analysis. J Stroke. 2023;25:223–232

20. Caplan LR. Lacunar infarction and small vessel disease: pathology and pathophysiology. J Stroke. 2015;17:2–6

21. Yoon Y, Lee DH, Kang DW, Kwon SU, Kim JS. Single subcortical infarction and atherosclerotic plaques in the middle cerebral artery: high-resolution magnetic resonance imaging findings. Stroke. 2013;44:2462–2467

22. Brown AJ, Teng Z, Evans PC, Gillard JH, Samady H, Bennett MR. Role of biomechanical forces in the natural history of coronary atherosclerosis. Nat Rev Cardiol. 2016;13:210–220

23. Kim BJ, Kim SM, Kang D, Kwon SU, Suh DC, Kim JS. Vascular tortuosity may be related to intracranial artery atherosclerosis. Int J Stroke. 2015;10:1081–1086

24. Li J, Bian Y, Wu F, Fan Z, Zhang C, Zhao X, et al. Association of morphology of lenticulostriate arteries and proximal plaque characteristics with single subcortical infarction: a whole-brain high-resolution vessel wall imaging study. J Am Heart Assoc. 2024;13

25. Ha SH, Jeong S, Park JY, Chang JY, Kang D, Kwon SU, et al. Association between arterial tortuosity and early neurological deterioration in lenticulostriate artery infarction. Sci Rep. 2023;13

26. Richardson PD, Davies MJ, Born GV. Influence of plaque configuration and stress distribution on fissuring of coronary atherosclerotic plaques. Lancet. 1989;2:941–944

27. Teng Z, Brown AJ, Calvert PA, Parker RA, Obaid DR, Huang Y, et al. Coronary plaque structural stress is associated with plaque composition and subtype and higher in acute coronary syndrome: the beacon i (biomechanical evaluation of atheromatous coronary arteries) study. Circ Cardiovasc Imaging. 2014;7:461–470

28. Nah HW, Kang DW, Kwon SU, Kim JS. Diversity of single small subcortical infarctions according to infarct location and parent artery disease: analysis of indicators for small vessel disease and atherosclerosis. Stroke. 2010;41:2822–2827

29. Warner JJ, Harrington RA, Sacco RL, Elkind M. Guidelines for the early management of patients with acute ischemic stroke: 2019 update to the 2018 guidelines for the early management of acute ischemic stroke. Stroke. 2019;50:3331–3332

30. Kleindorfer DO, Towfighi A, Chaturvedi S, Cockroft KM, Gutierrez J, Lombardi-Hill D, et al. 2021 guideline for the prevention of stroke in patients with stroke and transient ischemic attack: a guideline from the american heart association/american stroke association. Stroke. 2021;52:e364–e467

31. Gao S, Wang YJ, Xu AD, Li YS, Wang DZ. Chinese ischemic stroke subclassification. Front Neurol. 2011;2:6

32. Sedlaczek O, Caplan L, Hennerici M. Impaired washout--embolism and ischemic stroke: further examples and proof of concept. Cerebrovasc Dis. 2005;19:396–401

33. Quintero-Consuegra MD, Toscano JF, Babadjouni R, Nisson P, Kayyali MN, Chang D, et al. Encephaloduroarteriosynangiosis averts stroke in atherosclerotic patients with border-zone infarct: post hoc analysis from a performance criterion phase ii trial. Neurosurgery. 2021;88:E312–E318

34. Kim BJ, Yoon Y, Lee D, Kang D, Kwon SU, Kim JS. The shape of middle cerebral artery and plaque location: high-resolution mri finding. Int J Stroke. 2015;10:856–860

35. Kim BJ, Yang E, Kim N, Kim M, Kang D, Kwon SU, et al. Vascular tortuosity may be associated with cervical artery dissection. Stroke. 2016;47:2548–2552

36. Lau AY, Wong KS, Lev M, Furie K, Smith W, Kim AS. Burden of intracranial steno-occlusive lesions on initial computed tomography angiography predicts poor outcome in patients with acute stroke. Stroke. 2013;44:1310–1316

37. Bai X, Qu X, Nogueira RG, Chen W, Zhao H, Cao W, et al. Impact of immediate postrecanalization cooling on outcome in acute ischemic stroke patients with a large ischemic core: prospective cohort study. Int J Surg. 2024;110:2065–2070

38. Alawneh JA, Moustafa RR, Baron JC. Hemodynamic factors and perfusion abnormalities in early neurological deterioration. Stroke. 2009;40:e443–e450

39. Brisset M, Boutouyrie P, Pico F, Zhu Y, Zureik M, Schilling S, et al. Large-vessel correlates of cerebral small-vessel disease. Neurology. 2013;80:662–669

40. Shang K, Chen X, Cheng C, Luo X, Xu S, Wang W, et al. Arterial tortuosity and its correlation with white matter hyperintensities in acute ischemic stroke. Neural Plast. 2022;2022:1–10

41. Lin MP, Brott TG, Liebeskind DS, Meschia JF, Sam K, Gottesman RF. Collateral recruitment is impaired by cerebral small vessel disease. Stroke. 2020;51:1404–1410

42. Huang H, Virmani R, Younis H, Burke AP, Kamm RD, Lee RT. The impact of calcification on the biomechanical stability of atherosclerotic plaques. Circulation. 2001;103:1051–1056

43. Lee RT, Schoen FJ, Loree HM, Lark MW, Libby P. Circumferential stress and matrix metalloproteinase 1 in human coronary atherosclerosis. Implications for plaque rupture. Arterioscler Thromb Vasc Biol. 1996;16:1070–1073

44. Hallow KM, Taylor WR, Rachev A, Vito RP. Markers of inflammation collocate with increased wall stress in human coronary arterial plaque. Biomech Model Mechanobiol. 2009;8:473–486

45. Rocha M, Jovin TG. Fast versus slow progressors of infarct growth in large vessel occlusion stroke: clinical and research implications. Stroke. 2017;48:2621–2627

46. Yamauchi H, Higashi T, Kagawa S, Kishibe Y, Takahashi M. Impaired perfusion modifies the relationship between blood pressure and stroke risk in major cerebral artery disease. J Neurol Neurosurg Psychiatry. 2013;84:1226–1232

47. Sun X, Deng Y, Zhang Y, Yang M, Sun D, Nguyen TN, et al. Balloon angioplasty vs medical management for intracranial artery stenosis: the basis randomized clinical trial. JAMA. 2024;332:1059–1069

48. Jing J, Meng X, Zhao X, Liu L, Wang A, Pan Y, et al. Dual antiplatelet therapy in transient ischemic attack and minor stroke with different infarction patterns: subgroup analysis of the chance randomized clinical trial. JAMA Neurol. 2018;75:711–719

